# A Systematic Review of the Effects of Diet on the Gut Microbiota in Individuals at Risk for Colorectal Cancer

**DOI:** 10.1101/2025.04.14.25325698

**Authors:** Saurabh Pandey, Jenna Skidmore, Jaapna Dhillon

## Abstract

Colorectal cancer (CRC) poses a major public health concern, with emerging evidence highlighting the critical role of the gut microbiota in CRC progression. Considering that diet is a major modifiable factor influencing CRC risk, partly through its interaction with the gut microbiome, this systematic review evaluated the effects of dietary intervention RCTs, including supplements, functional foods, whole foods, and dietary patterns, on the gut microbiome in at-risk individuals (PROSPERO: CRD42024530038). A search of PubMed, Scopus, and Web of Science identified 4,746 records, with 20 additional records sourced manually. Following screening, six studies met inclusion criteria, focusing on microbial diversity, taxonomic composition, and metabolites. Findings demonstrate that whole food interventions like navy beans and functional foods like rice bran increased microbial diversity over 4-8 wk; with no effects of rice bran over 24 wk. Navy beans (8 wk) enriched beneficial taxa (*Faecalibacterium*, *Bifidobacterium*), while a shorter 4-wk intervention using bean powder showed no taxonomic shifts. Both forms of navy bean increased amino acid derivatives and anti-inflammatory phenolic metabolites. Rice bran increased *Lactobacillus* over 24 wk but showed differing effects on Firmicutes over short (2 wk) versus longer (24 wk) durations. Rice bran also increased SCFAs (acetate, propionate) over 2 wk and phenolic metabolites (e.g., diosmin) while reducing pro-carcinogenic byproducts (p-cresol sulfate, glycodeoxycholate) over 4 wk. β-glucan-enriched bread increased acetate over 12 wk but had minimal effects on microbial composition. Healthy Eating and Mediterranean dietary patterns did not alter taxonomic composition over 26 wk but reduced branched-chain bacterial fatty acids, indicating reduced proteolytic fermentation. This review underscores the potential of dietary strategies to modulate the gut microbiome in CRC-risk populations. However, limited RCTs and heterogeneity limit generalizability. Future research should conduct rigorous RCTs across the lifespan, using advanced microbiome and metabolite analyses and examining understudied dietary patterns to guide CRC prevention.

## Introduction

Colorectal cancer (CRC) is one of the most frequently diagnosed cancers globally and accounts for a considerable proportion of cancer-associated illness and death, creating a substantial public health burden (1). Recent projections indicate a dramatic rise in colorectal cancer incidence among young adults, with expected increases of up to 124% for those aged 20–34 and nearly 46% for individuals aged 35–49 by the year 2030 (2). Globally, CRC incidence is three to four times higher in developed countries than in developing regions, reflecting its association with socioeconomic development (3).

Diet plays a pivotal role in CRC risk influencing its development through interactions with the gut microbiome (4). Adherence to dietary patterns such as the Mediterranean diet, Healthy Eating Index-2005, or prudent dietary patterns, and other patterns rich in fruits, vegetables, and whole grains, is associated with reduced CRC risk (relative risk [RR] 0.45–0.90) (5).

Conversely, dietary patterns dominated by processed and red meats, refined foods, and Western diets are linked to significantly increased CRC risk (RR 1.18–11.7) (5). Similarly, cohort and case-control studies in young adults at risk for early-onset CRC show that diets high in sugar-sweetened beverages, deep-fried foods, and low in folate and fiber are associated with increased cancer risk, while fruit and vegetable-based diets rich in micronutrients are protective (6).

The gut microbiome mediates diet-related effects on CRC risk. Dysbiosis, or a disruption in microbial balance, contributes to chronic inflammation, genotoxic metabolite production, and immune dysregulation, all of which are implicated in CRC development (4,7,8). For instance, high fat diets have been shown to stimulate specific gut microbiota to produce carcinogenic secondary bile acids, which can promote CRC development (9). In contrast, short chain fatty acids (SCFAs), produced from the fermentation of dietary fibers, have demonstrated protective effects against CRC, suggesting that a high-fiber diet may mitigate cancer risk (9,10).

Recent advancements in metagenomics and metabolomics have further elucidated these diet-microbiome-CRC links. Meta-analyses of metagenomic datasets from various geographic regions have identified globally associated CRC microbes, including *Fusobacterium nucleatum*, *Peptostreptococcus stomatis*, and *Gemella morbillorum*, among others such as *Bacteroides fragilis*, *Parvimonas* spp., *Clostridium symbiosum*, and *Solobacterium moorei* (11). These microbial shifts are associated with CRC-enriched predicted metabolites such as amino acids, cadaverine, and creatine (11), as well as secondary bile acids (12). Additionally, functional analyses have highlighted CRC-enriched pathways, such as protein and mucin catabolism, alongside a depletion in carbohydrate degradation pathways (12). These findings underscore the complex interplay between dietary factors, gut microbiota, and CRC development.

Despite these advances, significant gaps remain in understanding how specific dietary interventions influence the gut microbiome in at-risk populations. Prior systematic reviews have largely focused on diet and microbiome outcomes in general populations, diet and CRC risk, or observational studies (6,13–22). This systematic review addresses this gap by synthesizing evidence from RCTs examining dietary interventions such as whole foods, enriched foods, supplements, and dietary patterns and their impact on microbial diversity, taxonomic composition, and metabolite production. By clarifying these relationships, this review aims to inform CRC prevention strategies, guide future research, and support evidence-based dietary recommendations for at-risk populations.

## Methods

The study protocol is registered with PROSPERO, the International Prospective Register of Systematic Reviews (CRD42024530038).

### Search strategy

A search strategy, developed in accordance with the Cochrane Handbook of Systematic Reviews (23), is detailed in **Supplementary Table S1**. The search was designed to identify dietary interventions examining the gut microbiota in individuals at risk for colorectal cancer. Primary searches were conducted on April 09, 2024 with no restrictions being imposed on the date of publication.

### Study selection

The search was conducted in three databases (PubMed, Scopus, and Web of Sciences) and the results were compiled in Zotero (version 6.0.23). To identify additional relevant studies, reference lists of related systematic reviews and meta-analyses were manually reviewed. Three authors (JD, SP, JS) systematically assessed the articles for inclusion. Discrepancies were resolved through a voting process. Details of the PICO framework can be found in **Supplementary Table S2**.

The study selection process is illustrated in the PRISMA (Preferred Reporting Items for Systematic Reviews and Meta-Analyses) flow diagram (24) (**Figure 1**). The exclusion process was conducted in four stages. In the first pass, studies were excluded if they were duplicates, involved children, pregnant individuals, non-human subjects, or in vitro experiments. Reviews, study protocols, non-research articles, genome analyses, text-mining, food nutrient analysis, food quality studies, and studies where the full text or abstract was not available in English were also excluded.

**Figure 1:**
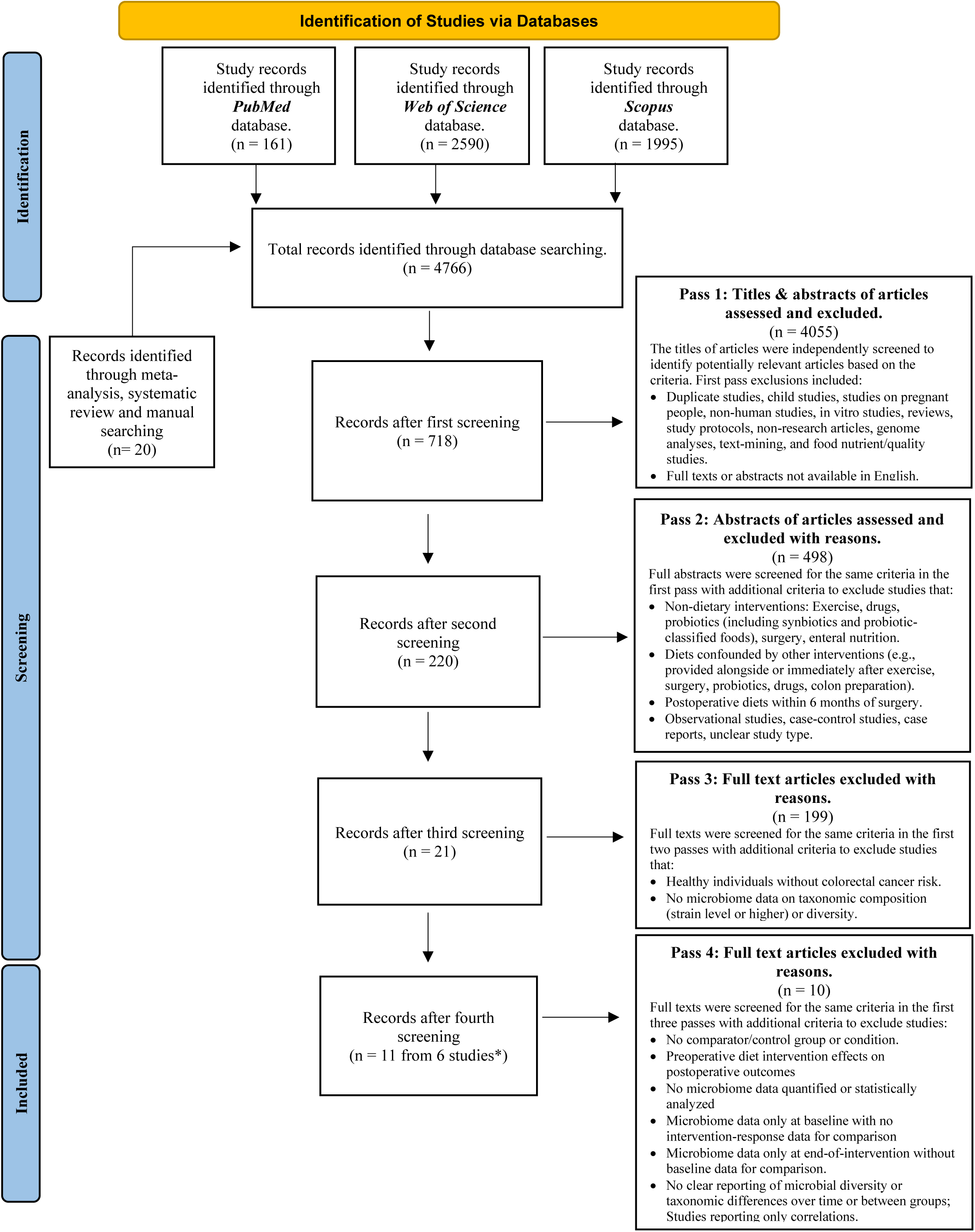
PRISMA Flow Diagram Summarizing the Study Selection Process. Two studies analyzed had multiple associated papers. *, 2 studies had multiple papers

In the second pass, exclusions included studies focusing on interventions other than diet, such as exercise, drugs, probiotics (including foods classified as probiotics by the study), synbiotics, surgery, or enteral nutrition. Dietary supplements described as drugs or registered with the FDA or international regulatory agencies as drugs were also excluded. Studies where dietary effects could not be isolated, such as those involving diets provided alongside or immediately after exercise, surgery, probiotics, drugs, or colon preparation interventions, were excluded. Diets provided postoperatively within six months of surgery, where surgical effects could confound outcomes, were also excluded. Observational studies, case-control studies, case reports, and studies unclear about whether they were observational or dietary interventions were removed.

In the third pass, studies were excluded if the population were healthy without colorectal cancer risk or if colorectal cancer risk was classified solely based on diet. Studies were also excluded if they did not provide microbiome data related to taxonomic composition (strain level or higher) or diversity. Additionally, studies presenting microbiome data only at the ASV/OTU level without aggregation to taxonomic composition were excluded.

In the fourth pass, exclusions were based on whether the study addressed the research question. Studies examining preoperative dietary interventions effects on postoperative outcomes were excluded. Studies were excluded if no microbiome data were quantified or statistically analyzed; if microbiome data were only presented at baseline with no intervention response data presented for comparison; or if microbiome data were only presented at the end of the intervention without baseline data for comparison. Additional exclusions included studies without a comparator or control group/condition, studies without clear reporting of differences in microbial diversity or taxonomic abundances over time or between groups. Studies that reported only correlations were also excluded. Papers replicating information already presented in another paper from the same study were not included in the review.

### Data extraction

Data collection tables were designed and finalized by three authors (JD, SP, JS). Data extraction was performed by SP and JS, with JD reviewing for accuracy. Study quality was assessed using the Oxford Centre for Evidence-Based Medicine Levels of Evidence (25), and risk of bias was evaluated using the Academy of Nutrition and Dietetics quality control checklists (26).

## Results

In total 4746 records were screened for inclusion from database search and 20 from meta-analyses, systematic reviews, and a manual review of reference lists. A total of 4762 records were excluded based on the established criteria (**Figure 1**). Six studies with dietary interventions that discussed microbiome outcomes in individuals at risk for CRC were included in the systematic review. Two studies had multiple publications discussing different microbiome outcomes (27–31) and are grouped together in **Table 1**.

**Table 1:**
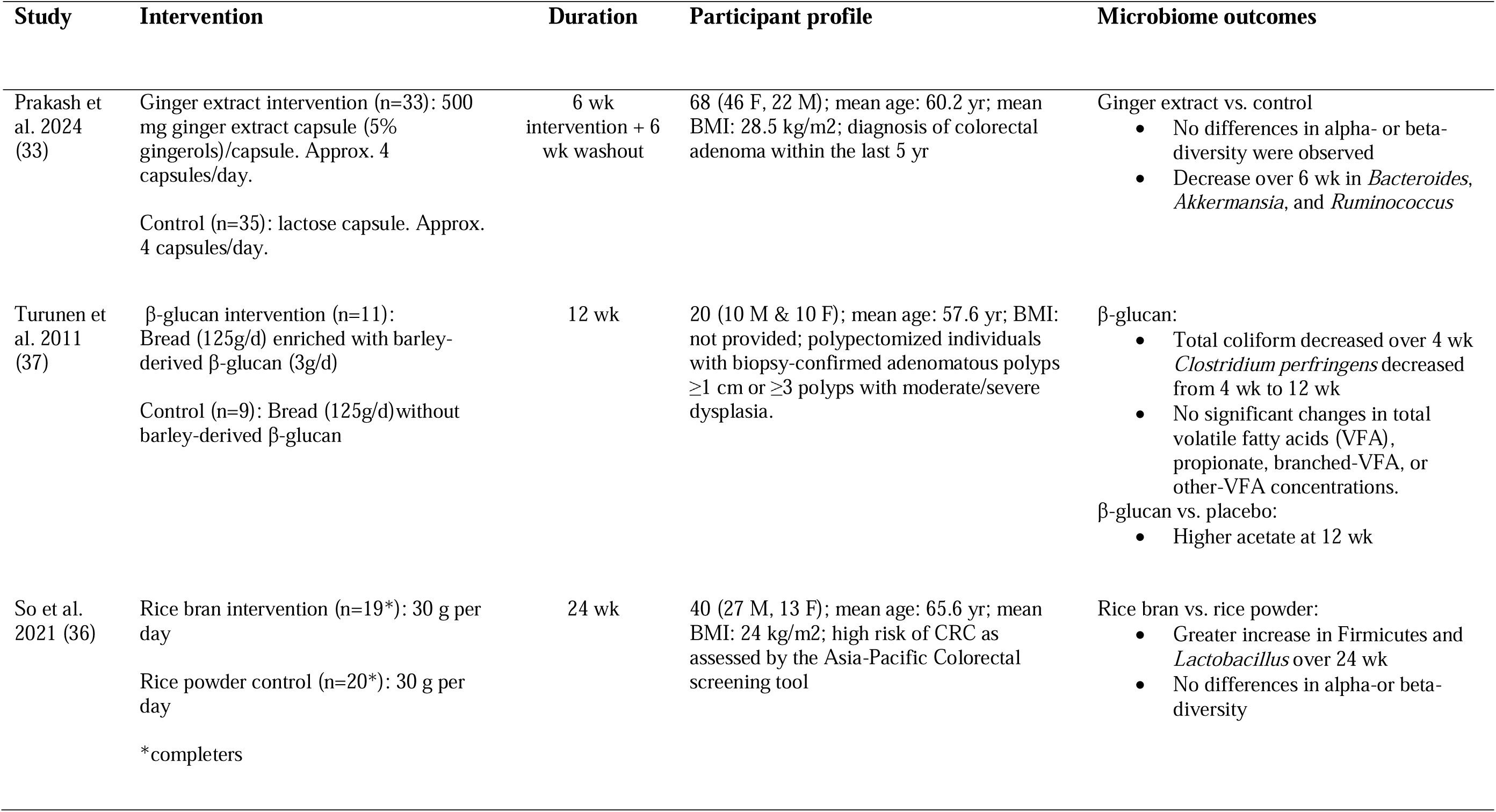
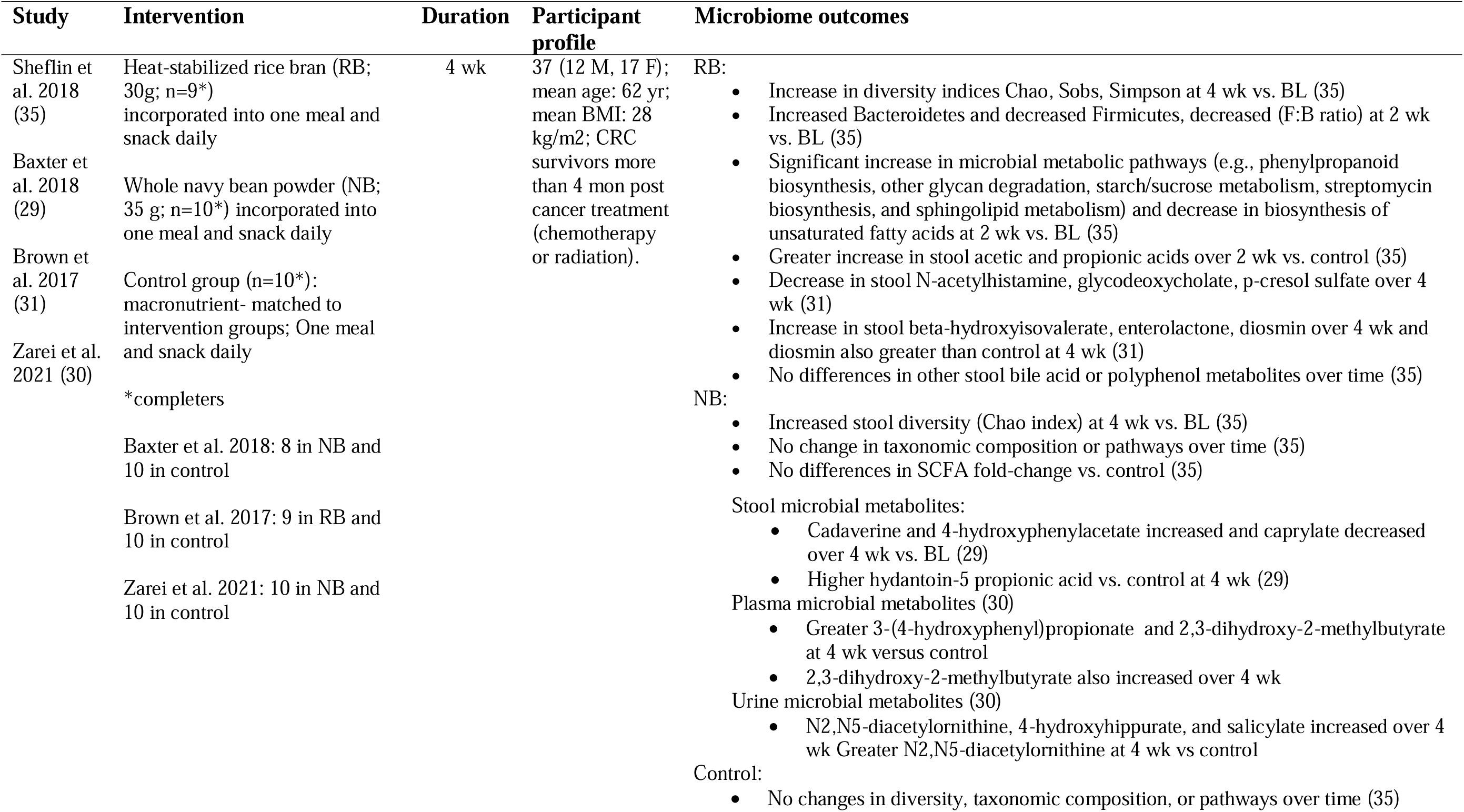
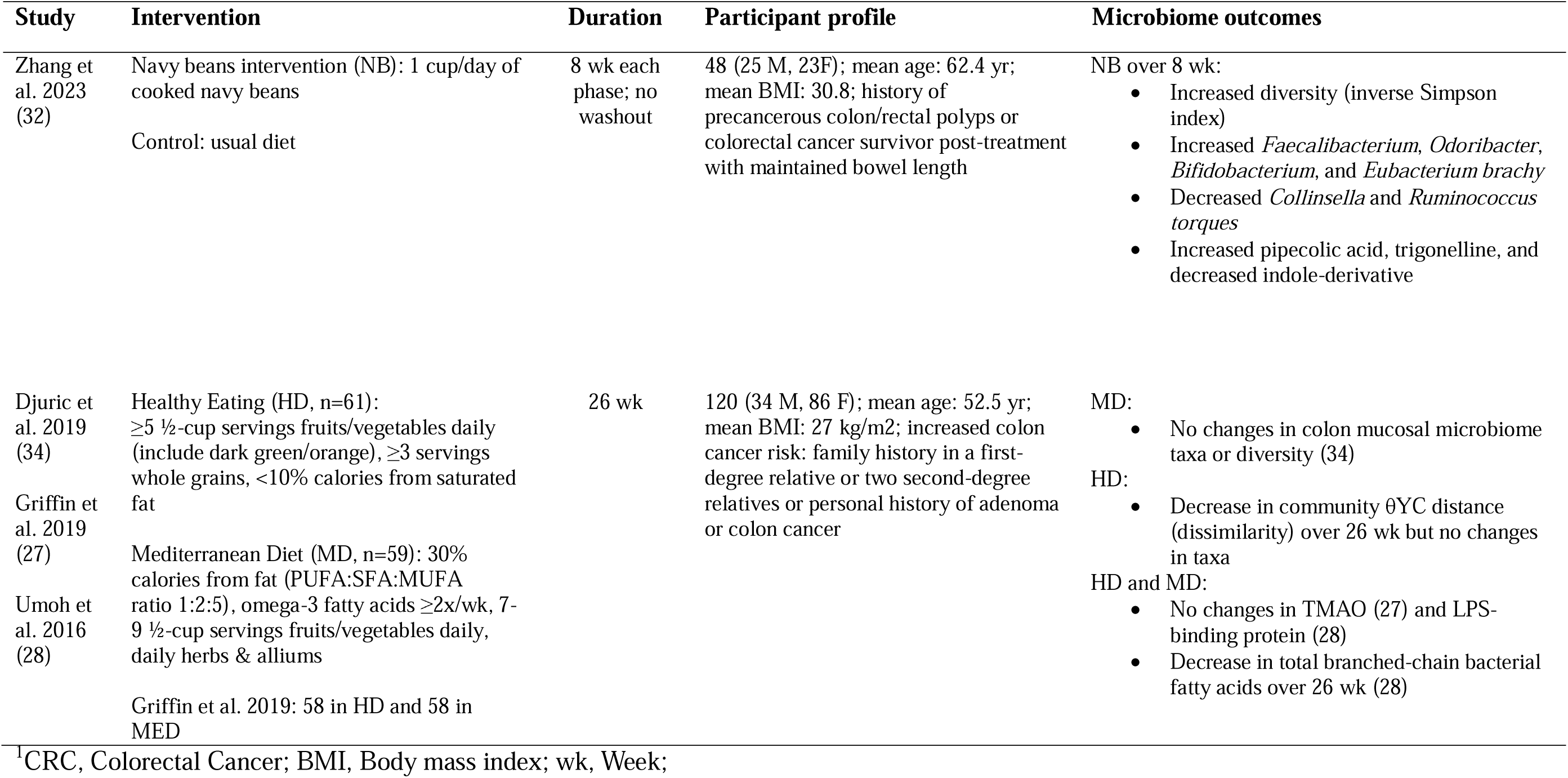
Characteristics of Studies Examining Dietary Interventions and Gut Microbiota in Individuals at Risk for Colorectal Cancer.

### Study design and duration

Four studies were conducted in the United States (32–35), 1 in China (36), 1 in Greece (37). Included studies varied in design and intervention duration tested (**Table 1**). Five studies (27–31,34–38) used parallel-arm randomized controlled trial (RCT) designs while one study (32) employed a randomized crossover design. Study durations ranged from 4 wk to 156 wk.

### Dietary interventions

The interventions studied were diverse ranging from dietary supplements such as ginger extract (33), functional foods such as rice bran (36) and β-glucan-enriched bread (37), whole foods such as navy beans (32), enriched meals and snacks containing rice bran or whole navy bean powder (35), and dietary patterns such as healthy eating or Mediterranean dietary patterns (34).

### Population characteristics

The studies examined diverse populations with varying CRC risks and treatment histories. Participants included individuals with adenomatous polyps (37), colorectal adenomas (33), and precancerous colon or rectal polyps (32). Other studies defined risk based on clinical tools such as the Asia-Pacific Colorectal Screening Tool (36) or family or personal history of adenomas or colon cancer (27,28,34). Several studies included CRC survivors more than four months post-treatment for chemotherapy or radiation (29–31,35). Participants across studies were predominantly middle-aged to older adults with mean ages ranging from the 50s to 70s and mean BMIs in the overweight and obesity range.

### Effects of dietary intervention on microbial diversity

Navy bean consumption increased stool microbial diversity, with significant increases in diversity indices such as the inverse Simpson index over 8 wk (32) and Chao index over 4 wk (35). Similarly, rice bran interventions showed an increase in Chao, Sobs, and Simpson diversity indices over 4 wk (35), but no differences in diversity over 24 wk (36). Dietary patterns such as Healthy Eating and Mediterranean diets had no significant impact on colon mucosal microbiome diversity. However, a decrease in community θYC distance, indicating reduced dissimilarity, was observed in the Healthy Eating group over 26 wk (34). Ginger extract supplement did not result in significant differences in alpha or beta diversity (33). The β-glucan intervention did not assess diversity indices (37).

### Effects of dietary intervention on microbial taxonomic composition

Navy bean consumption over 8 wk resulted in increased abundance of *Faecalibacterium*, *Odoribacter*, *Bifidobacterium*, and *Eubacterium brachy*, with decreases in *Collinsella* and *Ruminococcus torques* (32). However, in another intervention, whole navy bean powder showed no changes in taxonomic composition over 4 wk (35). In the same study, rice bran increased the abundance of Bacteroidetes while decreasing Firmicutes, leading to a reduction in the Firmicutes-to-Bacteroidetes ratio at 2 wk compared to baseline (35). In a 24 wk duration rice bran intervention, a greater increase in Firmicutes and *Lactobacillus* compared to rice powder was observed (36). The ginger extract intervention resulted in significant decreases in *Bacteroides*, *Akkermansia*, and *Ruminococcus* compared to placebo over 6 wk (33). The β-glucan intervention decreased total coliform abundance over 4 wk and *Clostridium perfringens* abundance from 4 to 12 wk (37). No significant changes in taxonomic composition were observed with Healthy Eating or Mediterranean diets in colon mucosal microbiota (34).

### Effects of dietary intervention on microbial-produced metabolites

Navy bean consumption over 8 wk increased pipecolic acid and trigonelline while decreasing an indole derivative (32). In the other navy bean intervention, stool cadaverine and 4-hydroxyphenylacetate increased while caprylate decreased over 4 wk (29). Additionally, stool hydantoin-5-propionic acid (29), plasma 3-(4-hydroxyphenyl)propionate, and plasma 2,3-dihydroxy-2-methylbutyrate were higher at 4 wk compared to control (30). Urine metabolites, including N2,N5-diacetylornithine, 4-hydroxyhippurate, and salicylate, increased over 4 wk (30). In the same study, rice bran increased stool acetic and propionic acids over 2 wk compared to control (35) and decreased stool N-acetylhistamine, glycodeoxycholate, and p-cresol sulfate over 4 wk (31). Additional increases in stool beta-hydroxyisovalerate, enterolactone, and diosmin were observed over 4 wk (31). The β-glucan intervention showed no significant changes in total volatile fatty acids (VFA), propionate, branched VFAs, or other-VFA concentrations. However, higher acetate concentrations were observed at 12 wk compared to placebo (37). A decrease in total branched-chain bacterial fatty acids (28) but no changes in trimethylamine-N-oxide (TMAO) (27) or lipopolysaccharide-binding protein (LPS-binding protein) (28) were observed in Healthy Eating and Mediterranean diets over 26 wk.

### Quality of assessment of studies

The characteristics of the included studies are summarized in **Table 2**. Assessment was done on all 6 studies including the multiple publications from 2 studies (27–31). According to the Oxford Centre for Evidence-Based Medicine levels of evidence (25), 3 articles (32,33,36) were classified as 1b, indicating high-quality individual RCTs, while 8 (27–31,34,35,37) was classified as 2b, indicating articles with less than 80% follow-up, participant retention, or participant data analyzed.

**Table 2:** Risk-of-Bias Analysis of Studies Examining Dietary Interventions and Gut Microbiota in Individuals at Risk for Colorectal Cancer.

| Study | Selection of participants free from bias? | Study with >80% follow up or participant data analyzed<sup>2</sup> | Standard/valid/reliable data collection procedures | QCC rating<sup>3</sup> | Evidence grade |
| --- | --- | --- | --- | --- | --- |
| Prakash et al. 2024 (33) | Yes | Yes | Yes | ⊕ | 1b |
| Turunen et al. 2011 (37) | Unclear | No | Yes | ∅ | 2b |
| So et al. 2021 (36) | Yes | Yes | Yes | ⊕ | 1b |
| Sheflin et al. 2018 (35) | Yes | No | Yes | ∅ | 2b |
| Baxter et al. 2018 (29) | Yes | No | Yes | ∅ | 2b |
| Brown et al. 2017 (31) | Yes | No | Yes | ∅ | 2b |
| Zarei et al. 2021 (30) | Yes | No | Yes | ∅ | 2b |
| Zhang et al. 2023 (32) | Yes | Yes | Yes | ⊕ | 1b |
| Djuric et al. 2019 (34) | Yes | No | Yes | ∅ | 2b |
| Griffin et al. 2019 (27) | Yes | No | Yes | ∅ | 2b |
| Umoh et al. 2016 (28) | Yes | No | Yes | ∅ | 2b |

<sup>1</sup>QCC, Quality Criteria Checklist.
<sup>2</sup>Follow-up here takes into account participant attrition or participants included in analysis.
<sup>3</sup>QCC rating (26): +, report has clearly addressed issues of inclusion/exclusion, bias, generalizability, and data collection and analysis; ∅, report is neither exceptionally strong nor exceptionally weak.
<sup>4</sup>Oxford Centre for Evidence-Based Medicine Levels of Evidence (25).

Using the Academy of Nutrition and Dietetics Quality Criteria Checklist for Primary Research risk-of-bias assessment (26), 3 articles (32,33,36) were rated positively (III), reflecting adequate consideration of bias, participant retention, and robustness, validity, and rigor in the intervention design, data collection, and analysis. Seven studies (27–31,34,35) were rated neutral (Ø), indicating they were neither exceptionally robust nor notably weak. One study (37) was rated negative, primarily because it was a pilot study that did not adequately address all validity criteria. Most studies in this review exhibited statistical limitations, particularly in assessing group-by-time interactions to analyze microbiome outcomes comprehensively.

Common issues included reliance on within-group changes without assessing intergroup differences over time in the same statistical model. Other issues pertained to lack of clarity in P-value reporting, and inconsistent application of false discovery rate (FDR) corrections for taxa-level analyses.

## Discussion

### Summary of evidence

This systematic review highlights the limited availability of RCT dietary interventions focusing on microbiome outcomes in individuals at risk for CRC. Existing evidence largely stems from observational studies examining associations between microbiome profiles and CRC risk, or diet and CRC risk (6,13–22). Additionally, many dietary intervention trials that were excluded focused on pre- or post-operative microbiome outcomes, recovery from surgery, or dietary effects combined with drugs or other therapies. Such studies, while valuable, do not provide direct insights into the potential of dietary interventions for modulating the microbiome for CRC prevention or recurrence in at-risk populations. In this context, the findings of this review shed light on how specific dietary interventions influence microbial diversity, taxonomic composition, and microbial-produced metabolites in CRC-risk individuals.

### Microbial diversity and taxonomic composition shifts

Functional compound interventions, such as ginger extract supplementation, showed no significant changes in stool diversity metrics, however, reductions in *Bacteroides*, *Akkermansia*, and *Ruminococcus* compared to placebo were observed (33). While Bacteroides includes both beneficial and pathogenic species (39), the reduction in *Akkermansia*, a bacterium involved in mucin degradation and gut barrier maintenance (40,41), and *Ruminococcus*, a genus of complex carbohydrate degraders (42), raises questions about whether these microbial shifts reflect unintended effects of ginger supplementation itself or are influenced by concurrent dietary changes during the intervention. The β-glucan-enriched bread intervention (37), derived from barley, demonstrated a fluctuating pattern regarding the abundance of pathogenic taxa *Clostridium perfringens* (43), and a decrease in total coliforms which seems to align with previous literature on oat or barley derived beta glucan (44,45), and could be a result of the limited ability of coliform strains to metabolize b-glucan (37).

Rice bran interventions (35,36) exhibited variable effects, with a 4-wk study reporting increased microbial diversity (35), while a 24-wk study showed no significant changes (36). Over 2 wk, rice bran increased abundance of *Bacteroidetes* and reduced the Firmicutes-to-Bacteroidetes ratio (35), which may reflect enhanced carbohydrate fermentation (46) and increased production of metabolites like propionate, known for their anti-inflammatory effects (47). Over 24 wk, rice bran was associated with an increase in *Lactobacillus* and Firmicutes compared to rice powder (36). *Lactobacillus*, consistent with rice bran’s prebiotic potential, contributes to gut homeostasis by reinforcing gut barrier integrity and preventing intestinal damage from microbial pathogens (48). However, the broader increase in Firmicutes remains challenging to interpret, as this phylum includes both beneficial and pro-inflammatory species (49,50), highlighting the importance of finer taxonomic and functional analyses.

Whole food interventions such as navy beans (32), demonstrated notable increases in microbial diversity, with significant improvements in alpha-diversity indices observed over 4 to 8 wk. Navy bean consumption enriched beneficial taxa like *Faecalibacterium* and *Bifidobacterium* (51–53), known for their anti-inflammatory properties and butyrate production (53–55), both critical in CRC prevention (56,57). Concurrently, pro-inflammatory taxa like *Collinsella* and *Ruminococcus torques* (58,59) decreased, further supporting the protective role of navy beans in modulating the gut microbiome. However, these effects were not replicated in an intervention using navy bean powder (35), which showed no significant taxonomic changes over a shorter duration. Processing, such as grinding beans into powder, may alter nutrient bioavailability (60,61) and microbial utilization patterns, but its impact on taxonomic composition remains unclear, emphasizing the need for comparative studies to evaluate the influence of food processing and baseline microbiota.

Dietary patterns such as Healthy Eating and Mediterranean diets (34) did not significantly alter taxonomic composition in colon mucosal microbiota, likely reflecting the inherent stability of the mucosal microbiota compared to the more dynamic stool microbiota. However, a reduction in θYC community dissimilarity within the Healthy Eating group over 26 wk suggests increased uniformity in microbial communities among participants. This stability in taxonomic composition may still coincide with functional or metabolite changes, highlighting potential shifts in microbial activity that contribute to CRC risk modulation.

Building on these structural changes in the microbiome, an examination of microbial metabolites shifts provides critical insights into the functional impacts of dietary interventions and their relevance to CRC prevention

### Microbial produced metabolites

Functional food interventions, such as rice bran and β-glucan-enriched bread, modulated microbial metabolism in ways potentially protective against CRC. Rice bran increased beneficial SCFAs including acetate and propionate (35), while simultaneously reducing harmful microbial byproducts such as glycodeoxycholate and p-cresol sulfate (31), suggesting suppression of tumor-promoting pathways (31,62–64,17,65,66). Similarly, β-glucan-enriched bread increased acetate levels (37), reinforcing the role of SCFAs in supporting epithelial repair, reducing inflammation, and maintaining gut integrity (67), key processes in CRC prevention (68,69).

Navy bean and rice bran interventions increased several polyphenol-derived metabolites, including enterolactone (29) and diosmin (31), both of which exhibit antioxidant and anti-inflammatory properties (70–77). Navy bean consumption also elevated salicylate (29), a plant-derived phenolic compound (78), known for its anti-inflammatory and anti-cancer effects (79). Notably, salicylate has been shown at pharmacological concentrations in vitro to reduce angiogenesis triggered by colon cancer cells (80). Another metabolite elevated by navy beans was 3-(4-hydroxyphenyl)propionate, a proanthocyanidin derivative (81) with demonstrated antioxidant and anti-inflammatory potential (82,83). In addition to phenol metabolites, navy bean interventions increased amino acid-related metabolites such as pipecolic acid (32) and cadaverine (29) which are associated with anti-inflammatory, anti-tumor effects (32,84,30), and gut barrier function (85,86). Interestingly, cadaverine is also elevated in CRC patients (87), suggesting its dual role as both a physiological modulator and a potential biomarker. Navy beans also increased plant alkaloid trigonelline (88,89), which has been linked to reduced CRC risk through suppression of Nrf2 activation, potentially limiting oxidative stress and tumor progression (90,91).

Dietary patterns, such as Healthy Eating and Mediterranean diets, shifted metabolic outputs away from proteolytic fermentation pathways, reducing metabolites like total branched-chain bacterial fatty acids. While some studies suggest a general protective effect of BCFAs on gut inflammation (92,93), elevated levels of fecal BCFAs have been found in CRC cohorts likely due to microbial fermentation of host-derived substrates such as sloughed intestinal cells (94). This shift in response to these dietary interventions may indicate less fermentation of host-derived substrates. **Figure 2** illustrates the effects of dietary interventions on microbial-derived metabolite changes with **Supplementary Table S3** providing additional context regarding the metabolite role and potential implications for CRC risk.

**Figure 2:**
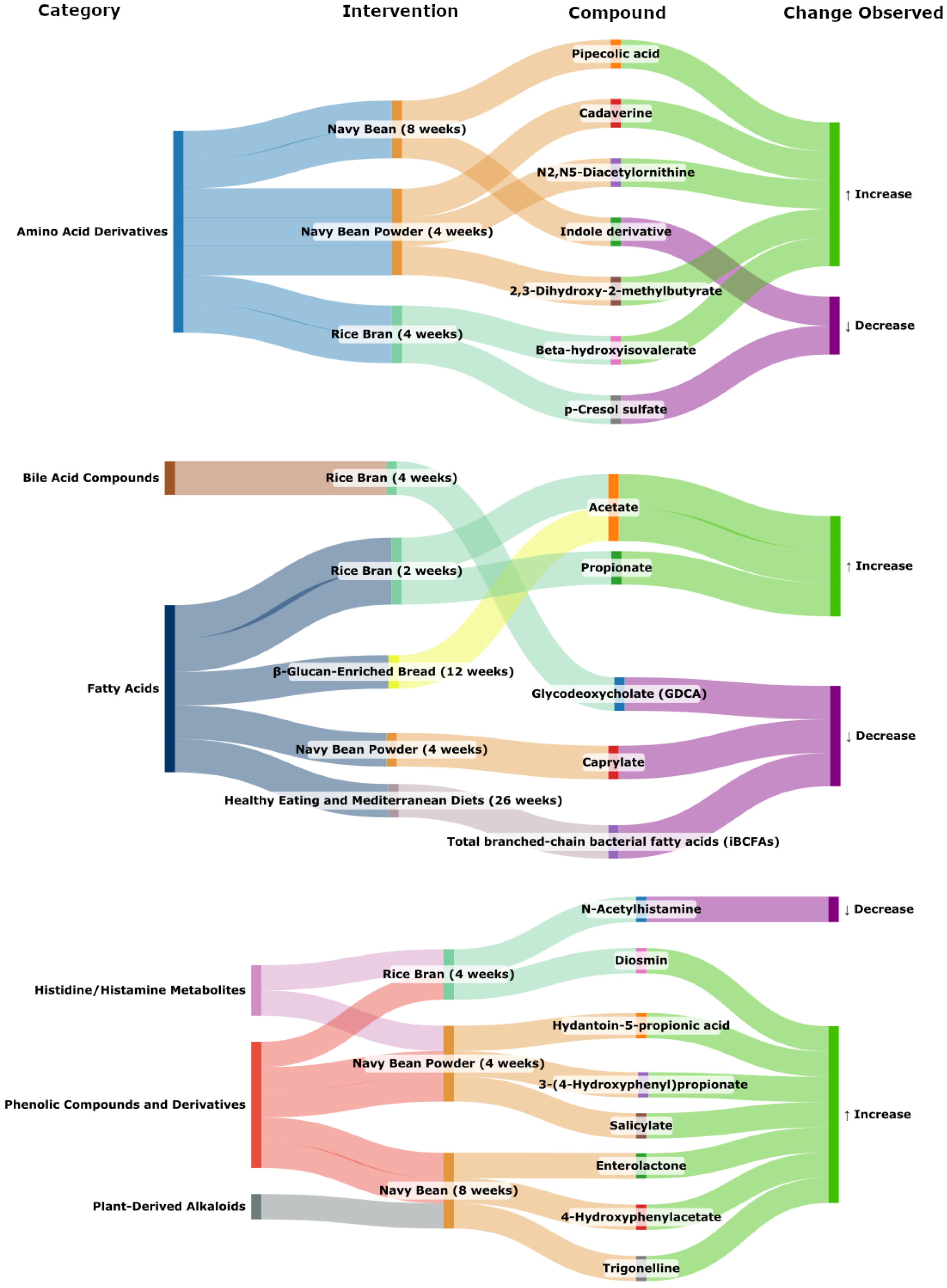
Sankey diagram illustrating the effects of dietary interventions on microbial-derived metabolite changes in individuals at risk for CRC. The first column categorizes metabolites based on their biochemical or functional classification and each class is assigned a unique color to indicate metabolite type. The second column lists dietary interventions, and the color serves as a linking factor between interventions and metabolites. The third column displays the specific microbial metabolites affected by these interventions and the color corresponds to the observed effect in response to the intervention, as indicated in the last column: green denotes increased metabolite levels, while purple indicates decreased levels.

The observed metabolite shifts underscore the microbiome’s critical role as a mediator between diet and host physiology, influencing colorectal cancer (CRC) risk. Microbial mechanisms contributing to CRC risk include the “driver-passenger" model theory which suggests that "driver" bacteria, initiate CRC by causing DNA damage, leading to the accumulation of mutations in colonic epithelial cells (15,95). These driver bacteria are eventually replaced by "passenger" bacteria, which thrive in the altered tumor microenvironment both promoting inflammation through activation of pathways such as NF-κB and Wnt/β-catenin, and tumor progression (15,95). Another theory purports that microbial dysbiosis can alter host energy metabolism by shifting from oxidative phosphorylation to glycolysis (Warburg effect), downregulating the tricarboxylic acid (TCA) cycle, and enhancing lactate production— hallmarks of cancer metabolism (21). Dietary interventions can modulate these microbiome-mediated pathways to reduce CRC risk. In vitro studies demonstrate that SCFAs can inhibit histone deacetylase (HDAC) activity, supporting tumor suppression pathways (96,97).

Resistant starch, which is readily metabolized by bacteria, and found in many foods, can influence CRC-related DNA methylation patterns, enhancing genome stability (e.g., GADD45A upregulation), decreasing cell cycle proliferation (e.g., CDK4 expression), and lowering crypt mitotic cell proportion in the upper half of the crypt in tumor tissue (13,98), contributing to anti-neoplastic effects. Additionally, polyphenols, either directly or through their microbial degradation byproducts, can reduce CRC risk by exerting antioxidant and anti-inflammatory effects, modulating signaling pathways, enhancing gut barrier integrity, promoting butyrate production, exhibiting selective antibacterial activity, and suppressing carcinogenic bacteria (22). This highlights the potential of dietary interventions to modulate these complex microbiome-mediated mechanisms.

### Strengths and Limitations

This systematic review’s strength lies in its exclusive focus on RCTs, providing a stronger quality of evidence on how dietary interventions influence the gut microbiome in individuals at risk for CRC. Many existing studies on diet and CRC have been prospective cohort or case-control studies, particularly in the context of microbiome outcomes. While such studies have been included in previous reviews (6,13–22), this review emphasizes the scarcity of RCTs, underscoring the need for more controlled studies, particularly in diverse populations. The review also provides a detailed analysis of microbial-derived metabolites, offering mechanistic insights into how dietary interventions may modulate CRC risk through metabolic pathways.

One limitation of this review is the inability to explore differences across the lifespan. Age-related changes in the gut microbiome such as reduced diversity and differences in taxonomic composition and functional capacity (99,100), may influence how individuals respond to dietary interventions. While older adults have traditionally been at higher CRC risk, recent trends indicate a plateau in this age group, whereas early-onset CRC is rising among younger adults (<50 years) (101,102). Moreover, while nutrient-rich dietary patterns consumed earlier in life may attenuate this risk (103,104), the lack of age-stratified RCTs prevented an in-depth analysis of these potentially critical life-stage differences. Additionally, the studies included in this review primarily focused on short- to medium-term dietary interventions, limiting insights into the long-term effects of diet on the microbiome and CRC risk. This constraint underscores the need for cautious interpretation of the findings.

### Implications of findings, research gaps, and future directions

A critical gap highlighted by this systematic review is the limited availability of RCTs examining the effects of dietary interventions on gut microbiome outcomes in individuals at risk for CRC. While the reviewed studies provide valuable insights into how specific foods and dietary patterns influence microbial diversity, taxonomic composition, and metabolite production, translating these findings into clinical strategies for CRC prevention requires deeper mechanistic insights.

Future studies should move beyond cataloging microbial composition and focus on how microbial functions influence CRC-related pathways, including inflammation, epithelial integrity, and immune modulation. One notable research gap is the underutilization of advanced methodologies to assess the functional capacity of the microbiome (105). While many studies have measured common microbial metabolites such as SCFAs and secondary bile acids, few have employed advanced functional analyses, meta-transcriptomics or metabolomics, to elucidate other microbial pathways involved in amino acid metabolism, polyphenol biotransformation, generation of bioactive metabolites, and their influence on host metabolic and immune responses. Incorporating these analyses into future dietary interventions could uncover novel mechanisms by which diet influences CRC risk through microbiome modulation The findings also underscore the need to examine whole dietary patterns rather than isolated food components. Much of the existing literature focuses on selected dietary components, such as fiber, without considering the broader dietary context that accounts for macronutrient and micronutrient interactions. For example, while fiber is widely recognized for its gut health benefits (106,107), its effects on the microbiome may be more pronounced within nutrient-rich diets that contain other healthy dietary components such as polyphenols (108). Similarly, fiber intake may counteract some of the adverse effects of high red meat consumption on CRC risk and inflammatory outcomes (20,109). It may not be individual dietary components alone but rather the overall dietary pattern that influences CRC risk. However, this nuanced approach, considering dietary patterns in RCTs to assess microbiome-related outcomes, remains underexplored in CRC prevention research.

Additionally, most studies have predominantly focused on Western diets and populations limiting the broader applicability of the findings. Expanding RCT research to include dietary patterns predominant in regions with low CRC risk such as Southern Asia, or Sub-Saharan Africa (110), could provide valuable insights into protective dietary components and distinct diet-microbiome interactions. This approach could help identify how interaction of specific dietary elements (e.g., high fiber content, unique polyphenols, or fermented foods) contribute to the reduced CRC risk observed in these populations.

To advance this field, future studies should prioritize robust trial designs with clear endpoints that also extend beyond microbiome outcomes to include host-relevant outcomes, such as gut barrier function, systemic inflammation, and specific cancer-related pathways. Utilizing insights from epidemiological and computational studies that highlight CRC-associated metabolic signatures could guide the testing of dietary interventions associated with specific microbial or metabolic pathways. Addressing these research gaps could pave the way for microbiome-targeted dietary strategies that are not only preventive but also tailored to individual risk profiles, potentially transforming CRC prevention and management strategies.

### Conclusion

This systematic review emphasizes the significant yet underexplored potential of dietary interventions to modulate gut microbiome pathways relevant to CRC prevention. While current evidence demonstrates that whole foods, nutrient-rich dietary patterns, and specific functional compounds can positively influence microbial diversity, composition, and metabolite production, substantial research gaps remain. Future studies must prioritize RCTs on dietary patterns in diverse populations and integrate advanced microbiome functional analyses to better delineate the diet-microbiome-CRC axis. Harnessing this knowledge could lead to tailored dietary strategies that effectively reduce CRC risk through microbiome modulation.

## Data Availability

All data produced in the present work are contained in the manuscript.

## Author contributions

The authors’ contributions were as follows—JD, SP, and JS: planned and developed the data collection tables; SP and JS: performed data extraction; JD: verified the accuracy of the extracted data; JD, SP, JS: conducted data analysis and prepared the manuscript; and all authors: reviewed, approved, and take full responsibility for the final manuscript.

## Conflict of interest

The authors report no conflicts of interest.

## Data Sharing

All data has been presented in the main manuscript and the supplementary file.

## Abbreviations used

CRC: Colorectal cancer
BMI: Body mass index
RCT: Randomized controlled trials
SCFA: Short chain fatty acids
wk: Week

## Notes

### Competing Interest Statement

The authors have declared no competing interest.

### Funding Statement

This study did not receive any funding

## References

1. Siegel RL, Giaquinto AN, Jemal A. Cancer statistics, 2024. CA A Cancer J Clinicians 2024;74:12–49.

2. Bailey CE, Hu C-Y, You YN, Bednarski BK, Rodriguez-Bigas MA, Skibber JM, Cantor SB, Chang GJ. Increasing Disparities in the Age-Related Incidences of Colon and Rectal Cancers in the United States, 1975-2010. JAMA Surg 2015;150:17.

3. Bray F, Ferlay J, Soerjomataram I, Siegel RL, Torre LA, Jemal A. Global cancer statistics 2018: GLOBOCAN estimates of incidence and mortality worldwide for 36 cancers in 185 countries. CA A Cancer J Clinicians 2018;68:394–424.

4. Siddiqui R, Boghossian A, Alharbi AM, Alfahemi H, Khan NA. The Pivotal Role of the Gut Microbiome in Colorectal Cancer. Biology 2022;11:1642.

5. Randi G, Edefonti V, Ferraroni M, Vecchia CL, Decarli A. Dietary Patterns and the Risk of Colorectal Cancer and Adenomas. Nutrition Reviews 2010;68:389–408.

6. Carroll KL, Frugé AD, Heslin MJ, Lipke EA, Greene MW. Diet as a Risk Factor for Early-Onset Colorectal Adenoma and Carcinoma: A Systematic Review. Front Nutr 2022;9:896330.

7. Artemev A, Naik S, Pougno A, Honnavar P, Shanbhag NM. The Association of Microbiome Dysbiosis With Colorectal Cancer. Cureus 14:e22156.

8. Dahmus JD, Kotler DL, Kastenberg DM, Kistler CA. The gut microbiome and colorectal cancer: a review of bacterial pathogenesis. J Gastrointest Oncol 2018;9:769–77.

9. Zeng H, Umar S, Rust B, Lazarova D, Bordonaro M. Secondary Bile Acids and Short Chain Fatty Acids in the Colon: A Focus on Colonic Microbiome, Cell Proliferation, Inflammation, and Cancer. Int J Mol Sci 2019;20:1214.

10. Chen Y, Yang Y, Gu J. Clinical Implications of the Associations Between Intestinal Microbiome and Colorectal Cancer Progression. Cancer Management and Research 2020;Volume 12:4117–28.

11. Avuthu N, Guda C. Meta-Analysis of Altered Gut Microbiota Reveals Microbial and Metabolic Biomarkers for Colorectal Cancer. Microbiol Spectr 2022;10:e0001322.

12. Wirbel J, Pyl PT, Kartal E, Zych K, Kashani A, Milanese A, Fleck JS, Voigt AY, Palleja A, Ponnudurai R, et al. Meta-analysis of fecal metagenomes reveals global microbial signatures that are specific for colorectal cancer. Nat Med 2019;25:679–89.

13. Clark MJ, Robien K, Slavin JL. Effect of prebiotics on biomarkers of colorectal cancer in humans: a systematic review. Nutrition Reviews 2012;70:436–43.

14. El Kinany K, Deoula M, Hatime Z, Bennani B, El Rhazi K. Dairy products and colorectal cancer in middle eastern and north African countries: a systematic review. BMC Cancer 2018;18:233.

15. Fratila TD, Ismaiel A, Dumitrascu DL. Microbiome modulation in the prevention and management of colorectal cancer: a systematic review of clinical interventions. Med Pharm Rep 2023;96:131–45.

16. Gianfredi V, Salvatori T, Villarini M, Moretti M, Nucci D, Realdon S. Is dietary fibre truly protective against colon cancer? A systematic review and meta-analysis. Int J Food Sci Nutr 2018;69:904–15.

17. Kim CE, Yoon LS, Michels KB, Tranfield W, Jacobs JP, May FP. The Impact of Prebiotic, Probiotic, and Synbiotic Supplements and Yogurt Consumption on the Risk of Colorectal Neoplasia among Adults: A Systematic Review. Nutrients 2022;14:4937.

18. Madrigal-Matute J, Bañón-Escandell S. Colorectal Cancer and Microbiota Modulation for Clinical Use. A Systematic Review. Nutr Cancer 2023;75:123–39.

19. Puzzono M, Mannucci A, Grannò S, Zuppardo RA, Galli A, Danese S, Cavestro GM. The Role of Diet and Lifestyle in Early-Onset Colorectal Cancer: A Systematic Review. Cancers (Basel) 2021;13:5933.

20. Turner ND, Lloyd SK. Association between red meat consumption and colon cancer: A systematic review of experimental results. Exp Biol Med (Maywood) 2017;242:813–39.

21. van Vorstenbosch R, Cheng HR, Jonkers D, Penders J, Schoon E, Masclee A, van Schooten F-J, Smolinska A, Mujagic Z. Systematic Review: Contribution of the Gut Microbiome to the Volatile Metabolic Fingerprint of Colorectal Neoplasia. Metabolites 2022;13:55.

22. Zhao Z, Feng Q, Yin Z, Shuang J, Bai B, Yu P, Guo M, Zhao Q. Red and processed meat consumption and colorectal cancer risk: a systematic review and meta-analysis. Oncotarget 2017;8:83306–14.

23. Cochrane Handbook for Systematic Reviews of Interventions [Internet]. [cited 2024 Dec 12]. Available from: https://training.cochrane.org/handbook

24. Page MJ, McKenzie JE, Bossuyt PM, Boutron I, Hoffmann TC, Mulrow CD, Shamseer L, Tetzlaff JM, Akl EA, Brennan SE, et al. The PRISMA 2020 statement: an updated guideline for reporting systematic reviews. BMJ 2021;372:n71.

25. Oxford Centre for Evidence-Based Medicine: Levels of Evidence (March 2009) — Centre for Evidence-Based Medicine (CEBM), University of Oxford [Internet]. [cited 2024 Dec 12]. Available from: https://www.cebm.ox.ac.uk/resources/levels-of-evidence/oxford-centre-for-evidence-based-medicine-levels-of-evidence-march-2009

26. EAL [Internet]. [cited 2024 Dec 12]. Available from: https://www.andeal.org/evidence-analysis-manual

27. Griffin LE, Djuric Z, Angiletta CJ, Mitchell CM, Baugh ME, Davy KP, Neilson AP. A Mediterranean diet does not alter plasma trimethylamine N-oxide concentrations in healthy adults at risk for colon cancer. Food Funct England; 2019;10:2138–47.

28. Umoh FI, Kato I, Ren J, Wachowiak PL, Ruffin MT 4th, Turgeon DK, Sen A, Brenner DE, Djuric Z. Markers of systemic exposures to products of intestinal bacteria in a dietary intervention study. Eur J Nutr Germany; 2016;55:793–8.

29. Baxter BA, Oppel RC, Ryan EP. Navy Beans Impact the Stool Metabolome and Metabolic Pathways for Colon Health in Cancer Survivors. Nutrients Switzerland; 2018;11.

30. Zarei I, Baxter BA, Oppel RC, Borresen EC, Brown RJ, Ryan EP. Plasma and Urine Metabolite Profiles Impacted by Increased Dietary Navy Bean Intake in Colorectal Cancer Survivors: A Randomized-Controlled Trial. Cancer Prev Res (Phila) United States; 2021;14:497–508.

31. Brown DG, Borresen EC, Brown RJ, Ryan EP. Heat-stabilised rice bran consumption by colorectal cancer survivors modulates stool metabolite profiles and metabolic networks: a randomised controlled trial. Br J Nutr England; 2017;117:1244–56.

32. Zhang X, Irajizad E, Hoffman KL, Fahrmann JF, Li F, Seo YD, Browman GJ, Dennison JB, Vykoukal J, Luna PN, et al. Modulating a prebiotic food source influences inflammation and immune-regulating gut microbes and metabolites: insights from the BE GONE trial. EBioMedicine Netherlands; 2023;98:104873.

33. Prakash A, Rubin N, Staley C, Onyeaghala G, Wen Y-F, Shaukat A, Milne G, Straka RJ, Church TR, Prizment A. Effect of ginger supplementation on the fecal microbiome in subjects with prior colorectal adenoma. Sci Rep England; 2024;14:2988.

34. Djuric Z, Bassis CM, Plegue MA, Ren J, Chan R, Sidahmed E, Turgeon DK, Ruffin MT, Kato I, Sen A. Colonic Mucosal Bacteria Are Associated with Inter-Individual Variability in Serum Carotenoid Concentrations. Journal of the Academy of Nutrition and Dietetics 2018;118:606–616.e3.

35. Sheflin AM, Borresen EC, Kirkwood JS, Boot CM, Whitney AK, Lu S, Brown RJ, Broeckling CD, Ryan EP, Weir TL. Dietary supplementation with rice bran or navy bean alters gut bacterial metabolism in colorectal cancer survivors. Mol Nutr Food Res Germany; 2017;61.

36. So WKW, Chan JYW, Law BMH, Choi KC, Ching JYL, Chan KL, Tang RSY, Chan CWH, Wu JCY, Tsui SKW. Effects of a Rice Bran Dietary Intervention on the Composition of the Intestinal Microbiota of Adults with a High Risk of Colorectal Cancer: A Pilot Randomised-Controlled Trial. Nutrients Switzerland; 2021;13.

37. Turunen K, Tsouvelakidou E, Nomikos T, Mountzouris K, Karamanolis D, Triantafillidis J, Kyriacou A. Impact of beta-glucan on the faecal microbiota of polypectomized patients: A pilot study. ANAEROBE 2011;17:403–6.

39. Zafar H, Saier MH. Gut Bacteroides species in health and disease. Gut Microbes 2021;13:1–20.

40. Elzinga J, Narimatsu Y, De Haan N, Clausen H, De Vos WM, Tytgat HLP. Binding of Akkermansia muciniphila to mucin is O-glycan specific. Nat Commun 2024;15:4582.

41. Mo C, Lou X, Xue J, Shi Z, Zhao Y, Wang F, Chen G. The influence of Akkermansia muciniphila on intestinal barrier function. Gut Pathog 2024;16:41.

42. La Reau AJ, Suen G. The Ruminococci: key symbionts of the gut ecosystem. J Microbiol 2018;56:199–208.

43. Mehdizadeh Gohari I, A. Navarro M, Li J, Shrestha A, Uzal F, A. McClane B. Pathogenicity and virulence of *Clostridium perfringens*. Virulence 2021;12:723–53.

44. Drzikova B, Dongowski G, Gebhardt E. Dietary fibre-rich oat-based products affect serum lipids, microbiota, formation of short-chain fatty acids and steroids in rats. British Journal of Nutrition 2005;94:1012–25.

45. Dongowski G, Huth M, Gebhardt E, Flamme W. Dietary Fiber-Rich Barley Products Beneficially Affect the Intestinal Tract of Rats. The Journal of Nutrition 2002;132:3704–14.

46. Flint HJ, Scott KP, Duncan SH, Louis P, Forano E. Microbial degradation of complex carbohydrates in the gut. Gut Microbes 2012;3:289–306.

47. Tedelind S, Westberg F, Kjerrulf M, Vidal A. Anti-inflammatory properties of the short-chain fatty acids acetate and propionate: A study with relevance to inflammatory bowel disease. WJG 2007;13:2826.

48. Dempsey E, Corr SC. Lactobacillus spp. for Gastrointestinal Health: Current and Future Perspectives. Front Immunol 2022;13:840245.

49. López-Bermudo L, Moreno-Chamba B, Salazar-Bermeo J, Hayward NJ, Morris A, Duncan GJ, Russell WR, Cárdenas A, Ortega Á, Escudero-López B, et al. Persimmon Fiber-Rich Ingredients Promote Anti-Inflammatory Responses and the Growth of Beneficial Anti-Inflammatory Firmicutes Species from the Human Colon. Nutrients 2024;16:2518.

50. Bolte LA, Vich Vila A, Imhann F, Collij V, Gacesa R, Peters V, Wijmenga C, Kurilshikov A, Campmans-Kuijpers MJE, Fu J, et al. Long-term dietary patterns are associated with pro-inflammatory and anti-inflammatory features of the gut microbiome. Gut 2021;70:1287–98.

51. Cronin M, Ventura M, Fitzgerald GF, Van Sinderen D. Progress in genomics, metabolism and biotechnology of bifidobacteria. International Journal of Food Microbiology 2011;149:4–18.

52. O’Callaghan A, Van Sinderen D. Bifidobacteria and Their Role as Members of the Human Gut Microbiota. Front Microbiol [Internet] 2016 [cited 2025 Feb 17];7. Available from: http://journal.frontiersin.org/Article/10.3389/fmicb.2016.00925/abstract

53. Sokol H, Pigneur B, Watterlot L, Lakhdari O, Bermúdez-Humarán LG, Gratadoux J-J, Blugeon S, Bridonneau C, Furet J-P, Corthier G, et al. *Faecalibacterium prausnitzii* is an anti-inflammatory commensal bacterium identified by gut microbiota analysis of Crohn disease patients. Proc Natl Acad Sci USA 2008;105:16731–6.

54. Rivière A, Gagnon M, Weckx S, Roy D, De Vuyst L. Mutual Cross-Feeding Interactions between Bifidobacterium longum subsp. longum NCC2705 and Eubacterium rectale ATCC 33656 Explain the Bifidogenic and Butyrogenic Effects of Arabinoxylan Oligosaccharides. Schloss PD, editor. Appl Environ Microbiol 2015;81:7767–81.

55. Imaoka A, Shima T, Kato K, Mizuno S, Uehara T, Matsumoto S, Setoyama H, Hara T, Umesaki Y. Anti-inflammatory activity of probiotic Bifidobacterium: Enhancement of IL-10 production in peripheral blood mononuclear cells from ulcerative colitis patients and inhibition of IL-8 secretion in HT-29 cells. WJG 2008;14:2511.

56. Faghfoori Z, Faghfoori MH, Saber A, Izadi A, Yari Khosroushahi A. Anticancer effects of bifidobacteria on colon cancer cell lines. Cancer Cell Int 2021;21:258.

57. Dikeocha IJ, Al-Kabsi AM, Chiu H-T, Alshawsh MA. Faecalibacterium prausnitzii Ameliorates Colorectal Tumorigenesis and Suppresses Proliferation of HCT116 Colorectal Cancer Cells. Biomedicines 2022;10:1128.

58. Astbury S, Atallah E, Vijay A, Aithal GP, Grove JI, Valdes AM. Lower gut microbiome diversity and higher abundance of proinflammatory genus *Collinsella* are associated with biopsy-proven nonalcoholic steatohepatitis. Gut Microbes 2020;11:569–80.

59. Schaus SR, Vasconcelos Pereira G, Luis AS, Madlambayan E, Terrapon N, Ostrowski MP, Jin C, Henrissat B, Hansson GC, Martens EC. *Ruminococcus torques* is a keystone degrader of intestinal mucin glycoprotein, releasing oligosaccharides used by *Bacteroides thetaiotaomicron*. Graf J, editor. mBio 2024;15:e00039–24.

60. Karam MC, Petit J, Zimmer D, Baudelaire Djantou E, Scher J. Effects of drying and grinding in production of fruit and vegetable powders: A review. Journal of Food Engineering 2016;188:32–49.

61. Cargo-Froom CL, Newkirk RW, Marinangeli CPF, Shoveller AK, Ai Y, Kiarie EG, Columbus DA. The effects of grinding and pelleting on nutrient composition of Canadian pulses. Can J Anim Sci 2022;102:457–72.

62. Rodríguez-Romero JDJ, Durán-Castañeda AC, Cárdenas-Castro AP, Sánchez-Burgos JA, Zamora-Gasga VM, Sáyago-Ayerdi SG. What we know about protein gut metabolites: Implications and insights for human health and diseases. Food Chemistry: X 2022;13:100195.

63. Al Hinai EA, Kullamethee P, Rowland IR, Swann J, Walton GE, Commane DM. Modelling the role of microbial p-cresol in colorectal genotoxicity. Gut Microbes 2019;10:398–411.

64. Sayin SI, Wahlström A, Felin J, Jäntti S, Marschall H-U, Bamberg K, Angelin B, Hyötyläinen T, Orešič M, Bäckhed F. Gut microbiota regulates bile acid metabolism by reducing the levels of tauro-beta-muricholic acid, a naturally occurring FXR antagonist. Cell Metab 2013;17:225–35.

65. Dai J, Wang, Hongxia, Dong, Ying, Zhang, Yinxin, and Wang J. Bile Acids Affect the Growth of Human Cholangiocarcinoma via NF-kB Pathway. Cancer Investigation Taylor & Francis; 2013;31:111–20.

66. Yuan Y, Yang C, Wang Y, Sun M, Bi C, Sun S, Sun G, Hao J, Li L, Shan C, et al. Functional metabolome profiling may improve individual outcomes in colorectal cancer management implementing concepts of predictive, preventive, and personalized medical approach. EPMA J 2022;13:39–55.

67. Liu P, Wang Y, Yang G, Zhang Q, Meng L, Xin Y, Jiang X. The role of short-chain fatty acids in intestinal barrier function, inflammation, oxidative stress, and colonic carcinogenesis. Pharmacological Research 2021;165:105420.

68. Mandle HB, Jenab M, Gunter MJ, Tjønneland A, Olsen A, Dahm CC, Zhang J, Sugier P-E, Rothwell J, Severi G, et al. Inflammation and gut barrier function-related genes and colorectal cancer risk in western European populations. Mutagenesis 2025;40:48–60.

69. Fan X, Jin Y, Chen G, Ma X, Zhang L. Gut Microbiota Dysbiosis Drives the Development of Colorectal Cancer. Digestion 2020;102:508–15.

70. Xiong X-Y, Hu X-J, Li Y, Liu C-M. Inhibitory Effects of Enterolactone on Growth and Metastasis in Human Breast Cancer. Nutrition and Cancer 2015;67:1326–34.

71. Mali AV, Padhye SB, Anant S, Hegde MV, Kadam SS. Anticancer and antimetastatic potential of enterolactone: Clinical, preclinical and mechanistic perspectives. European Journal of Pharmacology 2019;852:107–24.

72. Johnsen NF, Olsen A, Thomsen BLR, Christensen J, Egeberg R, Bach Knudsen KE, Loft S, Overvad K, Tjønneland A. Plasma enterolactone and risk of colon and rectal cancer in a case–cohort study of Danish men and women. Cancer Causes Control 2010;21:153–62.

73. Xie Y-Y, Yuan D, Yang J-Y, Wang L-H, Wu C-F. Cytotoxic activity of flavonoids from the flowers of *Chrysanthemum morifolium* on human colon cancer Colon205 cells. Journal of Asian Natural Products Research 2009;11:771–8.

74. Zheng Q, Hirose Y, Yoshimi N, Murakami A, Koshimizu K, Ohigashi H, Sakata K, Matsumoto Y, Sayama Y, Mori H. Further investigation of the modifying effect of various chemopreventive agents on apoptosis and cell proliferation in human colon cancer cells. Journal of Cancer Research and Clinical Oncology 2002;128:539–46.

75. Huwait E, Mobashir M. Potential and Therapeutic Roles of Diosmin in Human Diseases. Biomedicines 2022;10:1076.

76. Tanaka T, Makita H, Kawabata K, Mori H, Kakumoto M, Satoh K, Hara A, Sumida T, Tanaka T, Ogawa H. Chemoprevention of azoxymethane-induced rat colon carcinogenesis by the naturally occurring flavonoids, diosmin and hesperidin. Carcinogenesis 1997;18:957–65.

77. Kuntz S, Wenzel U, Daniel H. Comparative analysis of the effects of flavonoids on proliferation, cytotoxicity, and apoptosis in human colon cancer cell lines. Eur J Nutr 1999;38:133–42.

78. Hayat S, Ali B, Ahmad A. Salicylic Acid: Biosynthesis, Metabolism and Physiological Role in Plants. In: Hayat S, Ahmad A, editors. Salicylic Acid: A Plant Hormone. [Internet] Dordrecht: Springer Netherlands; 2007 [cited 2025 Apr 7]. p. 1–14. Available from: 10.1007/1-4020-5184-0_1

79. Shirakawa K, Wang L, Man N, Maksimoska J, Sorum AW, Lim HW, Lee IS, Shimazu T, Newman JC, Schröder S, et al. Salicylate, diflunisal and their metabolites inhibit CBP/p300 and exhibit anticancer activity. Shilatifard A, editor. eLife eLife Sciences Publications, Ltd; 2016;5:e11156.

80. Shtivelband MI, Juneja HS, Lee S, Wu KK. Aspirin and salicylate inhibit colon cancer medium- and VEGF-induced endothelial tube formation: correlation with suppression of cyclooxygenase-2 expression. Journal of Thrombosis and Haemostasis 2003;1:2225–33.

81. Ward NC, Croft KD, Puddey IB, Hodgson JM. Supplementation with Grape Seed Polyphenols Results in Increased Urinary Excretion of 3-Hydroxyphenylpropionic Acid, an Important Metabolite of Proanthocyanidins in Humans. J Agric Food Chem 2004;52:5545– 9.

82. Zhang Y-Y, Li X-L, Li T-Y, Li M-Y, Huang R-M, Li W, Yang R-L. 3-(4-Hydroxyphenyl)propionic acid, a major microbial metabolite of procyanidin A2, shows similar suppression of macrophage foam cell formation as its parent molecule. RSC Adv 2018;8:6242–50.

83. Kucukgul A, Nita EO. Revista Científica Facultad de Ciencias Veterinarias. Revista Científica de la Facultad de Ciencias Veterinarias Facultad de Ciencias Veterinarias. Universidad del Zulia; 2024;34:7.

84. Perera T, Young MR, Zhang Z, Murphy G, Colburn NH, Lanza E, Hartman TJ, Cross AJ, Bobe G. Identification and monitoring of metabolite markers of dry bean consumption in parallel human and mouse studies. Molecular Nutrition Food Res 2015;59:795–806.

85. Hanus M, Parada-Venegas D, Landskron G, Wielandt AM, Hurtado C, Alvarez K, Hermoso MA, López-Köstner F, De La Fuente M. Immune System, Microbiota, and Microbial Metabolites: The Unresolved Triad in Colorectal Cancer Microenvironment. Front Immunol 2021;12:612826.

86. Coni S, Di Magno L, Serrao SM, Kanamori Y, Agostinelli E, Canettieri G. Polyamine Metabolism as a Therapeutic Target in Hedgehog-Driven Basal Cell Carcinoma and Medulloblastoma. Cells 2019;8:150.

87. Yang Y, Misra BB, Liang L, Bi D, Weng W, Wu W, Cai S, Qin H, Goel A, Li X, et al. Integrated microbiome and metabolome analysis reveals a novel interplay between commensal bacteria and metabolites in colorectal cancer. Theranostics 2019;9:4101–14.

88. Zhou J, Chan L, Zhou S. Trigonelline: A Plant Alkaloid with Therapeutic Potential for Diabetes and Central Nervous System Disease. CMC 2012;19:3523–31.

89. Zia T, Hasnain SN, Hasan SK. Evaluation of the oral hypoglycaemic effect of Trigonella foenum-graecum L. (methi) in normal mice. Journal of Ethnopharmacology 2001;75:191– 5.

90. Panieri E, Buha A, Telkoparan-Akillilar P, Cevik D, Kouretas D, Veskoukis A, Skaperda Z, Tsatsakis A, Wallace D, Suzen S, et al. Potential Applications of NRF2 Modulators in Cancer Therapy. Antioxidants 2020;9:193.

91. Boettler U, Sommerfeld K, Volz N, Pahlke G, Teller N, Somoza V, Lang R, Hofmann T, Marko D. Coffee constituents as modulators of Nrf2 nuclear translocation and ARE (EpRE)-dependent gene expression. The Journal of Nutritional Biochemistry 2011;22:426– 40.

92. Nikolaki MD, Kasti AN, Katsas K, Petsis K, Lambrinou S, Patsalidou V, Stamatopoulou S, Karlatira K, Kapolos J, Papadimitriou K, et al. The Low-FODMAP Diet, IBS, and BCFAs: Exploring the Positive, Negative, and Less Desirable Aspects—A Literature Review. Microorganisms 2023;11:2387.

93. Yan Y, Wang Z, Greenwald J, Kothapalli KSD, Park HG, Liu R, Mendralla E, Lawrence P, Wang X, Brenna JT. BCFA suppresses LPS induced IL-8 mRNA expression in human intestinal epithelial cells. Prostaglandins, Leukotrienes and Essential Fatty Acids 2017;116:27–31.

94. Gall GL, Guttula K, Kellingray L, Tett AJ, Hoopen R ten, Kemsley EK, Savva GM, Ibrahim A, Narbad A. Metabolite quantification of faecal extracts from colorectal cancer patients and healthy controls. Oncotarget Impact Journals; 2018;9:33278–89.

95. Tjalsma H, Boleij A, Marchesi JR, Dutilh BE. A bacterial driver–passenger model for colorectal cancer: beyond the usual suspects. Nat Rev Microbiol Nature Publishing Group; 2012;10:575–82.

96. Chang PV, Hao L, Offermanns S, Medzhitov R. The microbial metabolite butyrate regulates intestinal macrophage function via histone deacetylase inhibition. Proceedings of the National Academy of Sciences Proceedings of the National Academy of Sciences; 2014;111:2247–52.

97. McBain JA, Eastman A, Nobel CS, Mueller GC. Apoptotic death in adenocarcinoma cell lines induced by butyrate and other histone deacetylase inhibitors. Biochemical Pharmacology 1997;53:1357–68.

98. Dronamraju SS, Coxhead JM, Kelly SB, Burn J, Mathers JC. Cell kinetics and gene expression changes in colorectal cancer patients given resistant starch: a randomised controlled trial. Gut 2009;58:413–20.

99. Badal VD, Vaccariello ED, Murray ER, Yu KE, Knight R, Jeste DV, Nguyen TT. The Gut Microbiome, Aging, and Longevity: A Systematic Review. Nutrients 2020;12:3759.

100. Leite G, Pimentel M, Barlow GM, Chang C, Hosseini A, Wang J, Parodi G, Sedighi R, Rezaie A, Mathur R. Age and the aging process significantly alter the small bowel microbiome. Cell Reports [Internet] Elsevier; 2021 [cited 2025 Mar 31];36. Available from: https://www.cell.com/cell-reports/abstract/S2211-1247(21)01219-5

101. Sung H, Siegel RL, Laversanne M, Jiang C, Morgan E, Zahwe M, Cao Y, Bray F, Jemal A. Colorectal cancer incidence trends in younger versus older adults: an analysis of population-based cancer registry data. The Lancet Oncology Elsevier; 2025;26:51–63.

102. Zaki TA, Singal AG, May FP, Murphy CC. Increasing Incidence Rates of Colorectal Cancer at Age 50–54 Years. Gastroenterology 2022;162:964–965.e3.

103. Chu AH, Lin K, Croker H, Kefyalew S, Becerra-Tomás N, Dossus L, González-Gil EM, Ahmadi N, Park Y, Krebs J, et al. Dietary patterns and colorectal cancer risk: Global Cancer Update Programme (CUP Global) systematic literature review. The American Journal of Clinical Nutrition [Internet] 2025 [cited 2025 Mar 31]; Available from: https://www.sciencedirect.com/science/article/pii/S0002916525000899

104. Kohler LN, Garcia DO, Harris RB, Oren E, Roe DJ, Jacobs ET. Adherence to Diet and Physical Activity Cancer Prevention Guidelines and Cancer Outcomes: A Systematic Review. Cancer Epidemiol Biomarkers Prev 2016;25:1018–28.

105. Mattes RD, Rowe SB, Ohlhorst SD, Brown AW, Hoffman DJ, Liska DJ, Feskens EJM, Dhillon J, Tucker KL, Epstein LH, et al. Valuing the Diversity of Research Methods to Advance Nutrition Science. Advances in Nutrition 2022;13:1324–93.

106. Fu J, Zheng Y, Gao Y, Xu W. Dietary Fiber Intake and Gut Microbiota in Human Health. Microorganisms 2022;10:2507.

107. Klurfeld DM, Davis CD, Karp RW, Allen-Vercoe E, Chang EB, Chassaing B, Fahey GC, Hamaker BR, Holscher HD, Lampe JW, et al. Considerations for best practices in studies of fiber or other dietary components and the intestinal microbiome. Am J Physiol Endocrinol Metab 2018;315:E1087–97.

108. Edwards CA, Havlik J, Cong W, Mullen W, Preston T, Morrison DJ, Combet E. Polyphenols and health: Interactions between fibre, plant polyphenols and the gut microbiota. Nutr Bull 2017;42:356–60.

109. Kopp TI, Vogel U, Tjonneland A, Andersen V. Meat and fiber intake and interaction with pattern recognition receptors (TLR1, TLR2, TLR4, and TLR10) in relation to colorectal cancer in a Danish prospective, case-cohort study. The American Journal of Clinical Nutrition Elsevier; 2018;107:465–79.

110. Vabi BW, Gibbs JF, Parker GS. Implications of the growing incidence of global colorectal cancer. J Gastrointest Oncol 2021;12:S387–98.

111. Liu Y, Hou Y, Wang G, Zheng X, Hao H. Gut Microbial Metabolites of Aromatic Amino Acids as Signals in Host-Microbe Interplay. Trends Endocrinol Metab 2020;31:818–34.

112. Aoki R, Aoki-Yoshida A, Suzuki C, Takayama Y. Indole-3-Pyruvic Acid, an Aryl Hydrocarbon Receptor Activator, Suppresses Experimental Colitis in Mice. The Journal of Immunology 2018;201:3683–93.

113. Alexeev EE, Lanis JM, Kao DJ, Campbell EL, Kelly CJ, Battista KD, Gerich ME, Jenkins BR, Walk ST, Kominsky DJ, et al. Microbiota-Derived Indole Metabolites Promote Human and Murine Intestinal Homeostasis through Regulation of Interleukin-10 Receptor. The American Journal of Pathology 2018;188:1183–94.

114. Wlodarska M, Luo C, Kolde R, d’Hennezel E, Annand JW, Heim CE, Krastel P, Schmitt EK, Omar AS, Creasey EA, et al. Indoleacrylic Acid Produced by Commensal Peptostreptococcus Species Suppresses Inflammation. Cell Host and Microbe 2017;22:25–37.e6.

115. Zarei I, Baxter BA, Oppel RC, Borresen EC, Brown RJ, Ryan EP. Plasma and Urine Metabolite Profiles Impacted by Increased Dietary Navy Bean Intake in Colorectal Cancer Survivors: A Randomized-Controlled Trial. Cancer Prevention Research 2021;14:497–508.

116. Tang X, Zhuang HJ, Yu H. Evaluation of the genetic architecture of human blood metabolites on sudden cardiac arrest: A Mendelian randomization analysis [Internet]. Research Square; 2024 [cited 2025 Apr 4]. Available from: https://www.researchsquare.com/article/rs-4011125/v1

117. Razavi AC, Bazzano LA, He J, Fernandez C, Whelton SP, Krousel Wood M, Li S, Nierenberg JL, Shi M, Li C, et al. Novel Findings From a Metabolomics Study of Left Ventricular Diastolic Function: The Bogalusa Heart Study. J Am Heart Assoc 2020;9:e015118.

118. Luo S, Surapaneni A, Zheng Z, Rhee EP, Coresh J, Hung AM, Nadkarni GN, Yu B, Boerwinkle E, Tin A, et al. NAT8 Variants, N-Acetylated Amino Acids, and Progression of CKD. Clin J Am Soc Nephrol 2020;16:37–47.

119. Hill EB, Baxter BA, Pfluger B, Slaughter CK, Beale M, Smith HV, Stromberg SS, Tipton M, Ibrahim H, Rao S, et al. Plasma, urine, and stool metabolites in response to dietary rice bran and navy bean supplementation in adults at high-risk for colorectal cancer. Front Gastroenterol [Internet] Frontiers; 2023 [cited 2025 Apr 4];2. Available from: https://www.frontiersin.org/journals/gastroenterology/articles/10.3389/fgstr.2023.1087056/full

120. Lee AJ, Beno DWA, Zhang X, Shapiro R, Mason M, Mason-Bright T, Surber B, Edens NK. A 14C-leucine absorption, distribution, metabolism and excretion (ADME) study in adult Sprague–Dawley rat reveals β-hydroxy-β-methylbutyrate as a metabolite. Amino Acids 2015;47:917–24.

121. Hague A, Elder DJ, Hicks DJ, Paraskeva C. Apoptosis in colorectal tumour cells: induction by the short chain fatty acids butyrate, propionate and acetate and by the bile salt deoxycholate. Int J Cancer 1995;60:400–6.

122. Nagao K, Yanagita T. Medium-chain fatty acids: Functional lipids for the prevention and treatment of the metabolic syndrome. Pharmacological Research 2010;61:208–12.

123. Narayanan A, Ananda Baskaran S, Amalaradjou MAR, Venkitanarayanan K. Anticarcinogenic Properties of Medium Chain Fatty Acids on Human Colorectal, Skin and Breast Cancer Cells in Vitro. Int J Mol Sci 2015;16:5014–27.

124. Iemoto T, Nishiumi S, Kobayashi T, Fujigaki S, Hamaguchi T, Kato K, Shoji H, Matsumura Y, Honda K, Yoshida M. Serum level of octanoic acid predicts the efficacy of chemotherapy for colorectal cancer. Oncology Letters Spandidos Publications; 2019;17:831–42.

125. Blachier F, Mariotti F, Huneau JF, Tomé D. Effects of amino acid-derived luminal metabolites on the colonic epithelium and physiopathological consequences. Amino Acids 2007;33:547–62.

126. Weber AM, Ibrahim H, Baxter BA, Kumar R, Maurya AK, Kumar D, Agarwal R, Raina K, Ryan EP. Integrated Microbiota and Metabolite Changes following Rice Bran Intake during Murine Inflammatory Colitis-Associated Colon Cancer and in Colorectal Cancer Survivors. Cancers (Basel) 2023;15:2231.

127. Acuña I, Ruiz A, Cerdó T, Cantarero S, López-Moreno A, Aguilera M, Campoy C, Suárez A. Rapid and simultaneous determination of histidine metabolism intermediates in human and mouse microbiota and biomatrices. BioFactors 2022;48:315–28.

128. KEGG COMPOUND: C05565 [Internet]. [cited 2025 Apr 4]. Available from: https://www.genome.jp/dbget-bin/www_bget?cpd:C05565

129. MetaCyc L-histidine degradation VI [Internet]. [cited 2025 Apr 4]. Available from: https://solcyc.solgenomics.net/META/NEW-IMAGE?detail-level=3&object=HISHP-PWY&type=PATHWAY&utm_source=chatgpt.com

130. Duran M, Bruinvis L, Wadman SK. Quantitative gas chromatographic determination of urinary hydantoin-5-propionic acid in patients with disorders of folate/vitamin B12 metabolism. Journal of Chromatography B: Biomedical Sciences and Applications 1986;381:401–5.

131. Khanal R, Howard LR, Prior RL. Urinary Excretion of Phenolic Acids in Rats Fed Cranberry, Blueberry, or Black Raspberry Powder. J Agric Food Chem American Chemical Society; 2014;62:3987–96.

132. Liu T, Chen Z, Sun L, Xiong L. Role of blood metabolites in mediating the effect of gut microbiota on chronic gastritis. Microbiology Spectrum American Society for Microbiology; 2024;12:e01490–24.

133. Liu Z, Xi R, Zhang Z, Li W, Liu Y, Jin F, Wang X. 4-hydroxyphenylacetic acid attenuated inflammation and edema via suppressing HIF-1α in seawater aspiration-induced lung injury in rats. Int J Mol Sci 2014;15:12861–84.

134. Zhao H, Jiang Z, Chang X, Xue H, Yahefu W, Zhang X. 4-Hydroxyphenylacetic Acid Prevents Acute APAP-Induced Liver Injury by Increasing Phase II and Antioxidant Enzymes in Mice. Front Pharmacol 2018;9:653.

135. Nguyen V, Taine EG, Meng D, Cui T, Tan W. Pharmacological Activities, Therapeutic Effects, and Mechanistic Actions of Trigonelline. IJMS 2024;25:3385.

