## Supplementary Tables S1 to S3 for "A Systematic Review of the Effects of Diet on the Gut Microbiota in Individuals at Risk for Colorectal Cancer"

**Supplementary Table S1**. Details of search methods

| **Database** | **Web Address** | **Keywords** |
| --- | --- | --- |
| PubMed | <http://www.ncbi.nlm.nih.gov/pubmed/> | (Diet OR nutrition OR nutrients OR food OR Supplement) AND (colorectal OR colon OR rectal) AND (cancer OR neoplasm OR adenocarcinoma OR metastasis OR tumor) AND (microbiome OR microbiota OR microflora OR microbiomes OR microbe OR microorganism OR microbiology OR microbial OR flora OR bacteria) |
| Web of Science | <https://www.webofscience.com/> | Topic -- ( diet OR nutrition OR nutrients OR food OR supplement ) AND ( colorectal OR colon OR rectal) AND (cancer OR neoplasm OR adenocarcinoma OR metastasis OR tumor)AND microbiome OR microbiota OR microflora OR microbiomes OR microbe OR microorganism OR microbiology OR microbial OR flora OR bacteria ) |
| SCOPUS | <https://www.scopus.com/> | TITLE-ABS-KEY ( ( ( diet OR nutrition OR nutrients OR food OR supplement ) AND ( colorectal OR colon OR rectal ) AND ( cancer OR neoplasm OR adenocarcinoma OR metastasis OR tumor ) AND ( microbiome OR microbiota OR microflora OR microbiomes OR microbe OR microorganism OR microbiology OR microbial OR flora OR bacteria ) ) ) |

The databases (PubMed, Web of Science, and Scopus) were searched on April 09, 2024 by JD, SP, and JS. Following the final searches, the protocol was registered. Articles were reviewed and screened by three authors (JD, SP, JS) based on established criteria.

**Supplementary Table S2**. PICO framework guiding the systematic review

| **Category** | **Inclusion Criteria** | **Exclusion Criteria** |
| --- | --- | --- |
| **Population** | Human adult populations (over 18 years old); individuals at risk of colorectal cancer. | Children (<18 years); pregnant populations; non-human studies; studies that classify CRC risk solely based on dietary history without clinical, genetic evidence, or family history. |
| **Intervention** | Dietary interventions including dietary patterns, specific food-based or nutrient-based components, or dietary supplements. | Non-dietary interventions: Exercise, drugs, probiotics (including synbiotics and foods classified as probiotics), surgery, or enteral nutrition  Diet effects cannot be isolated: diets provided alongside or immediately after exercise, surgery, probiotics, drugs, or colon preparation interventions  Diets initiated within 6 months after surgery where surgical recovery confounds dietary effects. |
| **Comparison** | Controls, placebos, or other dietary intervention groups. | No comparator/control groups |
| **Outcomes** | Primary outcomes: microbial diversity (alpha, beta), taxonomic composition (strain level or higher); Secondary outcomes: microbiome-derived metabolites. | Studies that provide only ASVs or OTUs without aggregating taxonomic composition; no clear reporting of differences in microbial diversity or taxonomic abundances over time or between groups. |

**Supplementary Table S3**: Classification of Microbial-Derived Metabolites and Compounds by Biochemical Origin and Their Potential Implications for CRC Risk

| **Compound Category** | **Intervention** | **Metabolite** | **Change Observed** | **Potential Role and CRC-Relevant Implications of Metabolite** |
| --- | --- | --- | --- | --- |
| **Amino Acid Derivatives** | Navy bean (8 wk ) (32) | Pipecolic acid  *Lysine catabolite* (32) | ↑ | Supports anti-inflammatory responses, anti-tumor and antibiotic properties potentially (30,32,84). |
|  | Navy bean (8 wk ) (32) | Indole derivative  *Byproduct of aromatic amino acid metabolism* (111) | ↓ | Indole derivatives have been found to enhance gut epithelial barrier function, activate anti-inflammatory IL-10 signaling pathways, and suppress experimental colitis (111–114)  CRC implications not clear. |
|  | Navy bean powder (4 wk ) (29) | Cadaverine  *Biogenic amine* | ↑ | Involved in gut barrier function and epithelial cell turnover (85,86).  Higher cadaverine levels observed in CRC vs. healthy controls; potential biomarker for CRC differentiation pending broader validation (87). |
|  | Navy bean powder (4 wk ) (30) | N2,N5-diacetylornithine  *Byproduct of urea cycle* (116,117) | ↑ | Associated with diastolic dysfunction, chronic kidney disease (CKD), and sudden cardiac arrest (116–118).  CRC implications not clear. |
|  | Navy bean powder (4 wk ) (30) | 2,3-dihydroxy-2-methylbutyrate  *Byproduct of branched chain amino acid metabolism* (119) | ↑ | Role and CRC implications not clear. |
|  | Rice bran (4 wk ) (31) | Beta-hydroxyisovalerate  *Byproduct of leucine metabolism* (120) | ↑ | CRC implications are unclear. |
|  | Rice bran (4 wk ) (31) | p-cresol sulfate  *Byproduct of aromatic amino acid metabolism* (111) | ↓ | P-cresol induces DNA damage in colonocytes in vitro (63). |
| **Bile Acid Compounds** | Rice bran (4 wk ) (31) | Glycodeoxycholate (GDCA)  *Secondary bile acid* (31) | ↓ | Promotes tumorigenesis in vivo and inflammation in vitro in cholangiocarcinoma  (65) .  Enhanced both proliferation and migration of CRC cells in vitro (66)**.**  Higher GDCA concentrations in CRC patients versus controls, with strong diagnostic performance for distinguishing CRC status (66) |
| **Fatty Acids** | β-glucan-enriched bread (12 wk) (37)  Rice bran (2 wk) (35) | Acetate (35,37)  Propionate (35)  *Short chain fatty acids* | ↑ | Strengthens gut barrier integrity, reduces inflammation, supports epithelial repair (67).  Induce apoptosis in colon cancer cell lines (121) . |
|  | Navy bean powder (4 wk ) (29) | Caprylate  *Medium-chain fatty acid* (122) | ↓ | Reduced cancer cell viability and inhibited CRC cell proliferation in a dose-dependent manner, accompanied by upregulation of apoptosis-related genes and downregulation of cell cycle regulatory genes (123).  Lower circulating levels of caprylic acid (octanoic acid) were associated with better CRC prognosis compared to higher levels ‑‑‑‑(124). |
|  | Healthy Eating and Mediterranean Diets (26 wk ) (28) | Total branched-chain bacterial fatty acids (iBCFAs)  *Byproducts of protein fermentation* (125) | ↓ | Protective role in gut inflammation (35,92).  Although higher concentrations observed in CRC cohorts compared to controls likely owing to microbial fermentation of host-derived substrates such as sloughed intestinal cells (94) |
| **Histidine/Histamine Metabolites** | Rice bran (4 wk ) (31) | N-acetylhistamine  *Byproduct of histamine metabolism* (126) | ↓ | Stimulates gastric secretion (126,127).  CRC implications not clear. |
|  | Navy bean powder (4 wk ) (29) | Hydantoin-5-propionic acid    *Linked to histidine metabolism* (128–130) | ↑ | Role and CRC implications not clear. |
| **Phenolic Compounds and Derivatives** | Navy bean (8 wk ) (29) | Enterolactone  *Lignan metabolite* (71) | ↑ | In vivo and in vitro studies support its anticancer and antimetastatic properties for CRC (71).  Clinically, enterolactone is associated with reduced cancer risk (71). |
|  | Navy bean (8 wk ) (29) | 4-hydroxyphenylacetate  *Phenolic compound* (131) | ↑ | Potential anti-inflammatory and anti-oxidant properties (132–134).  Specific role in CRC implications not clear. |
|  | Navy bean powder (4 wk ) (30) | 3-(4-hydroxyphenyl)propionate  *Proanthocyanidin metabolite* (81) | ↑ | Counters oxidative stress and inflammation (82,83).  Role in CRC implications is unclear. |
|  | Navy bean powder (4 wk ) (29) | Salicylate  *Plant-derived phenolic derivative* (78) | ↑ | Associated with anti-inflammatory and anti-cancer effects (79).  At pharmacological concentrations in vitro shown to reduce angiogenesis triggered by colon cancer cells (80). |
|  | Rice bran (4 wk ) (31) | Diosmin  *Flavonoid* (75) | ↑ | Exhibit anti-oxidant and anti-cancer properties (75).  Anti-proliferative effects in human colon cancer cell lines and chemopreventive activity against colon carcinogenesis in animal models (76,77). |
| **Plant-Derived Alkaloids** | Navy bean (8 wk ) (32) | Trigonelline  *Plant alkaloid* (88,89) | ↑ | Supports antioxidant and anti-inflammatory properties (135).  May reduce CRC risk by suppressing Nrf2 activation, potentially limiting oxidative stress and tumor progression (90,91). |

Metabolites/compounds may fall into overlapping categories; classifications were selected based on biochemical context and relevance to the reviewed interventions.
